# Latent tuberculosis infection and incident cardiovascular and non-cardiovascular death

**DOI:** 10.1101/2024.03.11.24304070

**Authors:** Ita M Magodoro, Katalina A Wilkinson, Brian L Claggett, Aloice Aluoch, Mark J Siedner, Mpiko Ntsekhe, Ntobeko AB Ntusi, John M Nyirenda, Robert J Wilkinson

## Abstract

Active tuberculosis may heighten the risk of incident cardiovascular morbidity and premature mortality, whereas whether latent TB infection (LTBI) recapitulates these adverse outcomes is unclear. We evaluated the effect of LTBI on all-cause and cardiovascular-specific death among US adults who underwent tuberculin skin testing in 1999-2000 and were followed up to December 31^st^, 2019. We also examined the impact of co-occuring traditional risk factors on these outcomes. Adjustments were made for socio-economic and demographic factors. LTBI was defined as tuberculin skin induration ≥10mm, and cause of death as cardiovascular if from heart or cerebrovascular diseases, and non-cardiovascular if otherwise. LTBI was associated with increased of overall and non-cardiovascular specific death but not cardiovascular-specific death. Risk of death was highest when LTBI was comorbid LTBI with diabetes. LTBI may increase risk of death by mechanisms other than progression to active TB disease.

## Introduction

Infection with *Mycobacterium tuberculosis (M.tb)* is characterized by an immunopathological spectrum (1), (2), (3), (4), (5). Clinically overt or active TB disease is often associated with a prolonged systemic inflammatory response with symptomatic tissue destruction and remodeling (6). In pulmonary tuberculosis (PTB), the commonest form of active *M.tb* infection, this manifests as parenchymal damage with impairment of ventilation and perfusion, and increased odds of long-term respiratory morbidity and mortality (7), (8). Because of its profound systemic perturbations, *M.tb* infection may also result in damage to tissues and organ systems beyond its primary site (9), (10). Indeed, a recent systematic review summarizing available evidence indicates that a history of active TB independently confers an increased risk [relative risk 1.51 (95% CI: 1.16–1.97)] for subsequent cardiovascular disease (CVD)(11). Similarly, other lines of evidence have also noted association between prior tuberculosis and new onset diabetes mellitus(12), lung cancer (13), (14), Parkinson’s disease (15) and persistent neurological deficits (16), among other health impairments. In this regard, it remains to be ascertained whether latent TB infection (LTBI), at the opposite end of the immuno-pathological spectrum, may also influence these adverse outcomes, including cardiovascular morbidity and mortality (17), (18), (19).

Whilst ostensibly healthy, it may be that tuberculosis sensitized persons who have ongoing bacillary replication experience subclinical immune activation and systemic pathology leading non-specifically to future morbidity, including CVD (20), (18). A sparse and heterogenous literature reports, for example, increased odds of both prevalent and incident CVD in LTBI (21), (22), (23), (24), (25), (26). Further complicating this is the overlap of latent *M.tb* infection with increasing traditional risk factors like obesity, smoking and hypertension (27), (28). Their co-occurrence likely worsens cardiovascular outcomes due to synergistic immunopathology (27). It is known, for example, that the chronic cellular and cytokine immune responses induced by *M.tb* (20) overlap with those mediating cardiovascular degeneration driven by traditional risk factors in the general population (29). However, these relationship remains understudied notwithstanding the twin facts that a quarter of the global population has evidence of LTBI and that CVD incidence and prevalence are not adequately accounted for by traditional risk factors, especially in the world’s disproportionately affected regions. Using a well characterized US prospective cohort, we evaluated the long-term cardiovascular consequences of LTBI, and its impact on overall survival. We also explored how pre-specified co-occuring traditional risk factors might impact health outcomes associated with latent *M.tb* infection.

## Methods

### Data Sources

Data for this study are derived from the 1999-2000 National Health and Nutrition Examination Survey (NHANES) linked to the National Death Index (NDI) with mortality follow-up through to December 31^st^, 2019. The NHANES are biennial cross-sectional surveys of the health status of the non-institutionalized US population. Each survey is designed to be nationally representative of the US demographic and socio-economic composition through multistage probability cluster sampling. Participant data are collected through standardized questionnaires, physical examinations, and laboratory testing of blood samples. The NDI, on the other hand, is a centralized database of individual-level records of date-of-death and underlying cause-of-death records for the entire US and its territories. Causes of death in the NDI are defined according to the World Health Organization (WHO) 10^th^ revision of the International Statistical Classification of Diseases, Injuries, and Causes of Death (ICD-10) guidelines. The study procedures, data collection and quality control methods for the NHANES and NDI, including their linkage, are fully described elsewhere (30). All data used in this study are publicly accessible in a de-identified format, negating the need for prior institutional review board approval.

### Analytic Sample Selection

All participants in the 1999-2000 NHANES aged at least 18 years were eligible for inclusion in the present analysis. Participants were excluded if they had any prior diagnoses of cardiovascular disease (heart failure, coronary artery disease, stroke, myocardial infarction) and/or had missing data on latent TB infection (LTBI) status, mortality outcomes and other key study covariates. The latter included participant demographic and socio-economic indicators, biophysical measurements and traditional risk factors.

### Study Measures

#### Latent tuberculosis infection

LTBI status was ascertained by tuberculin skin testing (TST) using tuberculin-purified protein derivative (PPD) product, Tubersol® (Sanofi, Bridgewater, NJ). Skin induration was measured according to standardized procedures by trained technicians 48-72 hours after placement of PPD on the volar surface of the forearm. Induration ≥10mm was considered indicative of LTBI.(31) Chest radiographs were not collected nor were data on current tuberculosis symptoms and BCG vaccination status.

#### Mortality and time-to-death

We categorized underlying causes of death using WHO ICD-10 codes into (i) cardiovascular if including heart and cerebrovascular diseases; and (ii) non-cardiovascular if including any of malignant neoplasms, chronic lower respiratory diseases, unintentional injuries, dementias, diabetes mellitus, infections and parasitic diseases, urinary system diseases, and all other residual causes. The ICD-10 codes used to categorize the causes of death are listed in **Table E1** in the **Online Data Supplement**. Participants were followed from time of baseline interview in 1999/2000, to either date of death or the end of the study period (December 31^st^, 2019), whichever came first.

#### Traditional risk factors and sociodemographic covariates

We defined diabetes mellitus as any current use of clinician-prescribed insulin and/or oral hypoglycemic medicines and/or HbA1c ≥6.5%; and hypertension as any current use of clinician-prescribed anti-hypertensive medicines and/or systolic blood pressure (SBP) ≥140 mmHg and/or diastolic blood pressure (DBP) ≥90 mmHg. Participants were defined as smokers if they had smoked at least 100 cigarettes in their lifetime and reported any smoking in the last 30 days. Non-smokers were those who either never smoked more than 100 cigarettes in their lifetime or had smoked ≥100 cigarettes in their lifetime but reported no smoking in the preceding 30 days. We used body mass index (BMI) to classify participants as overweight/obese if BMI ≥25 kg/m^2^, and under-/normal weight if BMI <25kg/m^2^.

Participants described their race as any one of Hispanic, non-Hispanic white, non-Hispanic black, and non-Hispanic Asian/Other, and their country of birth as any one of US, Mexico or elsewhere. Their socioeconomic status was measured by family income-to-poverty ratio (PIR) and educational attainment, the latter graded as “less than high school”, “high school diploma”, and “college degree or higher”. Health insurance was current if a participant had coverage in the preceding 12 months. We also extracted information on current use of clinician-prescribed statins, oral anticoagulants, and/or aspirin. Glomerular filtration rate (eGFR), a proxy for kidney health, was estimated from race, sex, and serum creatinine level using the CKD-EPI (Chronic Kidney Disease Epidemiology Collaboration) equation.(32) Less than 0.6% of the participants tested positive for HIV antibodies, and thus we did not consider it further. Lastly, we estimated 30-years risk for atherosclerotic cardiovascular disease (ASCVD) using Framingham risk equations (33).

## Statistical Analysis

### Matching and covariate selection

To enhance the between-group comparability and thus attenuate the risk of biased or confounded estimates, we matched participants with LTBI with peers without TB infection using the propensity score (PS) matching weight approach (34), (35). Propensity scores were estimated using multivariable logistic regression models, with LTBI status as the dependent variable, and age, sex, race, country of birth, education attained, family PIR, health insurance status, BMI, eGFR, cigarette smoking, diabetes and hypertension status and current use of statins, oral anticoagulants, and aspirin as the predictors. These covariates were selected, based on current literature, because they were either potential confounders of the relationship between LTBI and cardiovascular/all-cause mortality or were independent risk factors for cardiovascular mortality, or both.

Matching was done 1:1 with nearest neighbor within a specified caliper distance of 0.2. Between-group post-match balance was assessed by estimating the standardized mean difference (SMD) for each of the baseline covariates. An SMD < 0.01 indicates no residual bias, while an SMD <0.10 indicates inconsequential residual bias.(34) Analyses using the unmatched sample were considered *unadjusted,* while those using the PS weight matched sample were considered *adjusted.* Descriptive statistics of study participants are presented, according to variable scale, as mean (SD), median (25^th^, 75^th^ percentile) or number (%).

### Time-to-event analysis

Our primary approach to survival analysis was to estimate restricted mean survival time (RMST) and restricted mean time lost (RMTL) (36). We modelled Cox proportional hazards, as outlined below, for our sensitivity analysis as well as to detect moderated effects (37), (38). RMST is defined as the amount of event-free survival time experienced over a specified time period, while RMTL is the amount of time lost due to an event within a specific period. The RMST is derived from the area under the Kaplan-Meier (KM) curve whereas the RMTL is the area under the cause-specific cumulative incidence function (CIF). We plotted (i) KM curves for all-cause death, and (ii) CIFs for cardiovascular and non-cardiovascular specific death (considered competing events) stratified by LTBI status in both unmatched (*unadjusted*) and matched (*adjusted*) samples. From these, we estimated RMST and RMTL at 18 years for each TB exposure group, as well as their differences between the TB negative versus LTBI groups. Eighteen (18) years was chosen because it was the minimum of the longest follow-up time of the two TB sensitization groups.

### Evaluating moderator effects of traditional risk factors

To assess whether and how the effect of LTBI on the risk of all-cause death was impacted by baseline traditional risk factors, the same RMST models were extended across pre-specified comorbid domains of diabetes, hypertension, obesity and cigarette smoking subgroups. Statistical significance for testing these potential moderator effects was set at Bonferroni-corrected two-sided α<0.025. This step was accompanied by formal tests for interaction between TB and the recognized risk factors of interest using Cox proportional hazards models (37), (38).

### Sensitivity analysis

Sensitivity analyses entailed three approaches. We estimated RMST for all-cause death using TST cut-off values of ≥15mm and ≥5mm to define LTBI (31), (39). Next, we repeated the survival analysis (for all-cause death) modelling Cox proportional hazards and determining whether the estimated hazard ratios (HR) were consistent with the RMST estimates. The proportional hazards assumption was tested by visual inspection of the KM curves and by the Grambsch and Therneau’s proposed test. The survival curves crossed, and we also found a violation for the exposure variable, diabetes (p=0.002). The latter warrants stratifying the Cox models on diabetes, which coincided with our assessment of moderated effects (outlined above). Thus, we only reported the RMST estimates using a TST cut-off value of ≥15mm as our sensitivity analysis. Our rationale was to test if and how our findings were sensitive to differences in diagnostic threshold for LTBI.

Analyses were conducted using R, version 3.6.3 (R Foundation for Statistical Computing, Vienna, Austria), and Stata version 17.0 (StataCorp, College Station, TX, USA). Except for Bonferroni corrected testing, where indicated, all probability values were 2-sided, with p-values <0.05 considered indicative of statistical significance.

## Results

### Baseline characteristics of matched sample

A total of 3,880 participants were included in the analytic sample (**Figure 1**). PS match weighting achieved a balance of baseline covariates (**Table 1**) and yielded an effective sample size of 604. The mean (SD) age at cohort entry was 50.2 (17.1) years for those with LTBI and 50.0 (20.2) years for uninfected controls. Most of the participants in the matched sample were male and four in ten were of Mexican birth. Participants had comparatively low socio-economic status. The highest educational attainment for approximately 67% of them was less than high school, while median family PIR was 1.5. A family PIR threshold <1.3 is considered living in poverty. On the other hand, most participants (approximately 70%) had health insurance coverage at time of cohort entry.

**Figure 1.**
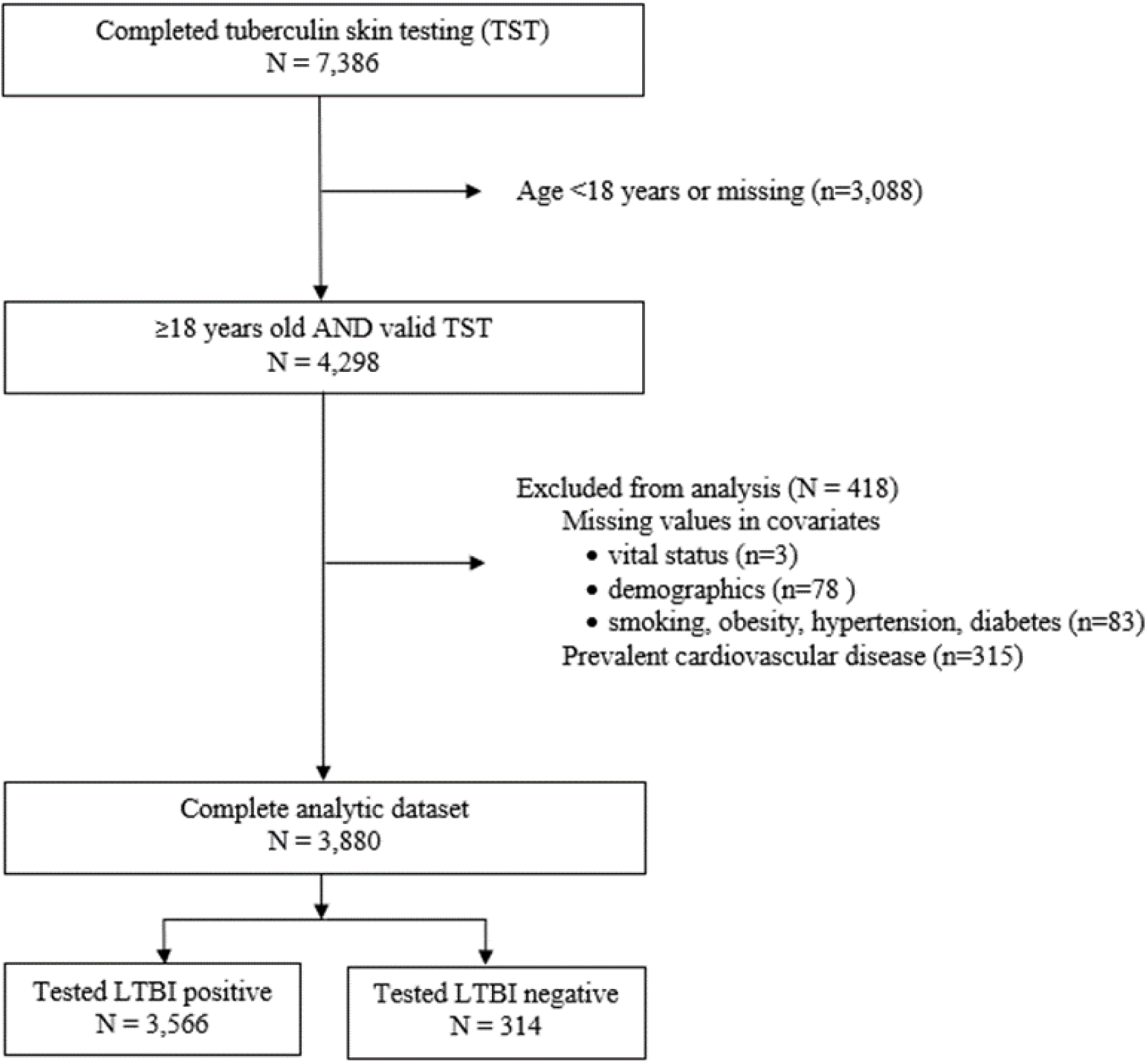
Flow chart of participant inclusion in final analytic sample using US NHANES 1999-2000

**Table 1.**
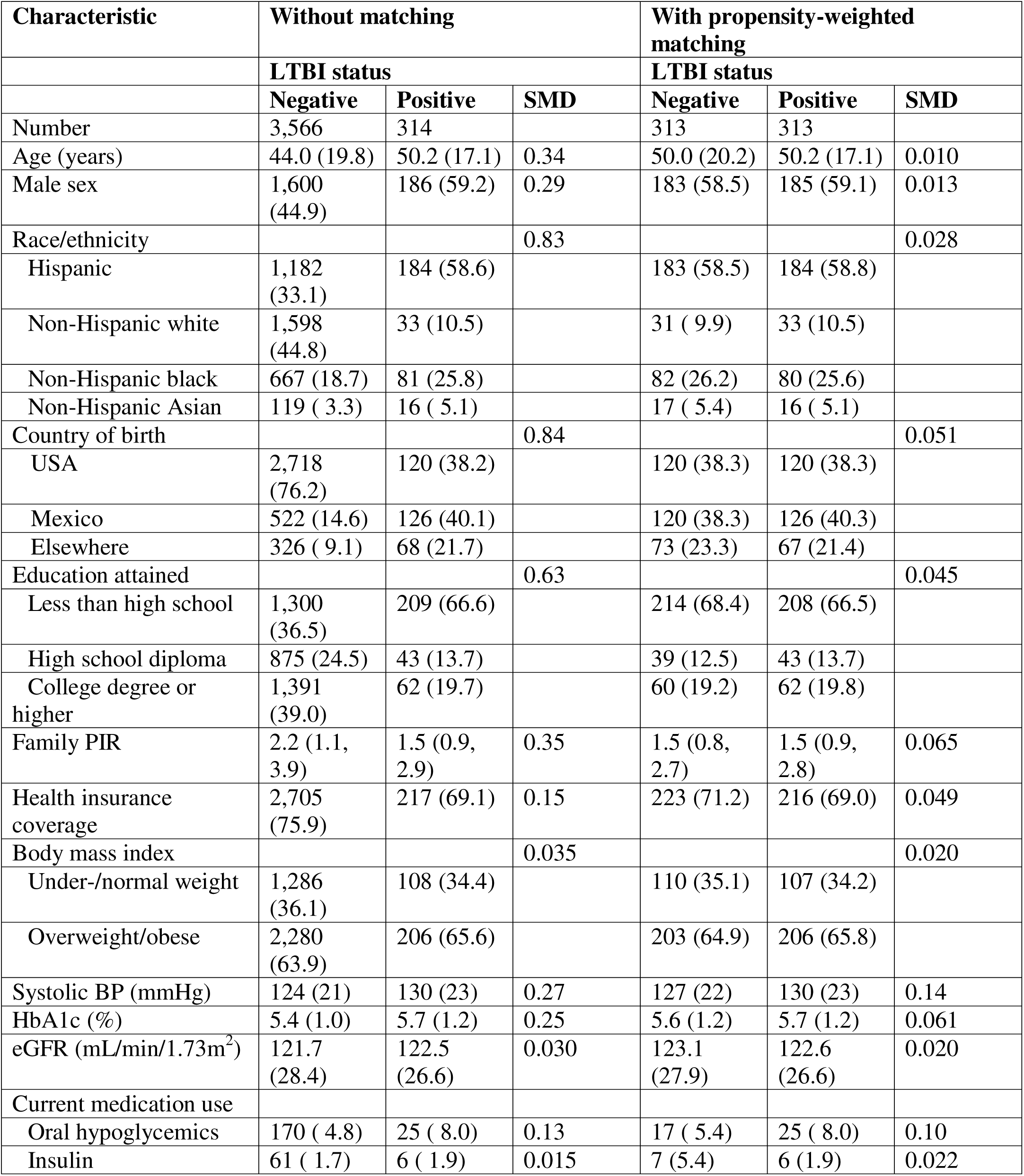
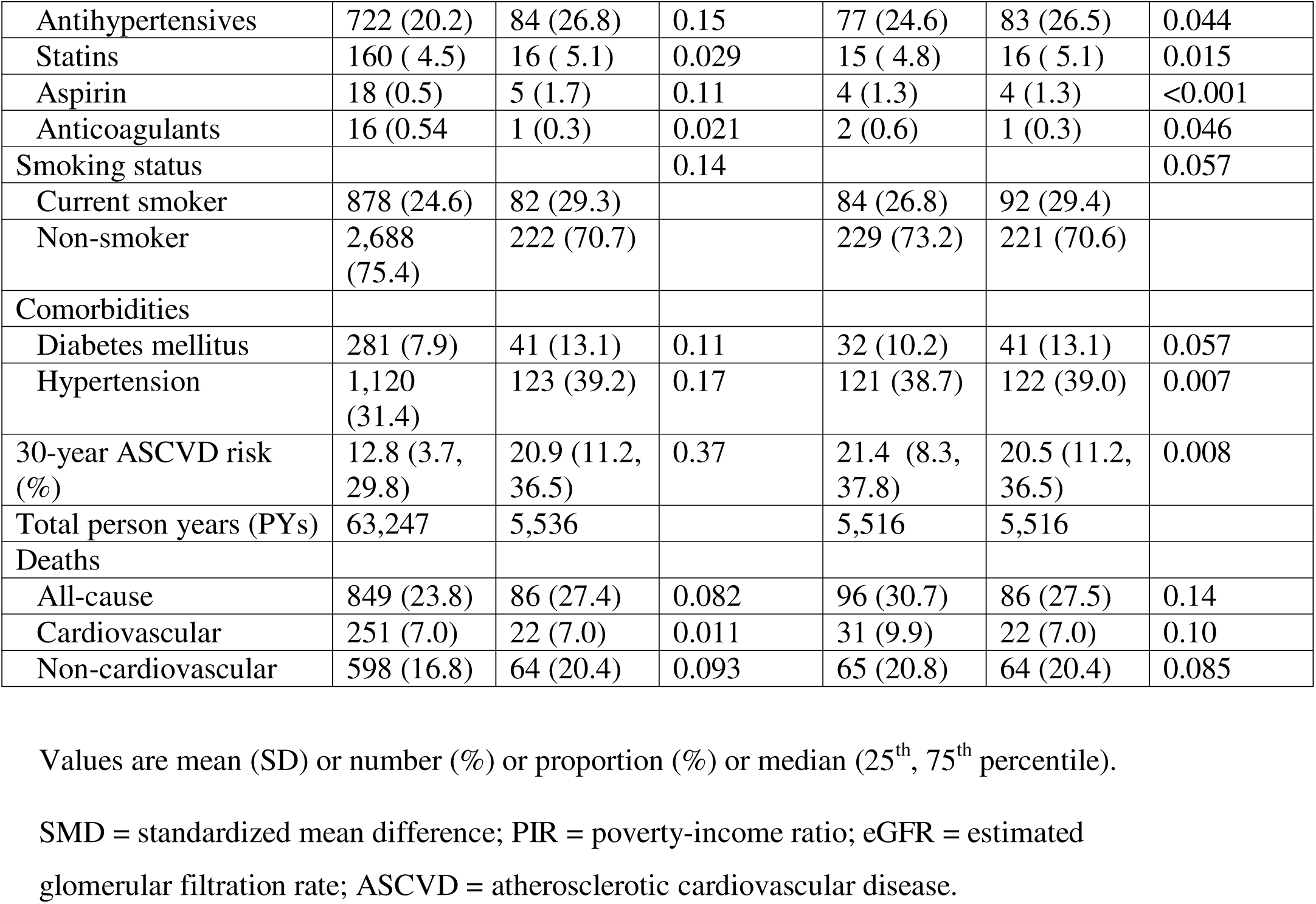
Participants’ characteristics stratified by latent tuberculosis infection status before and after propensity-weighted matching, US NHANES 1999-2000.

### Distribution of traditional risk factors

Before matching, participants with LTBI had worse cardiometabolic profiles compared to their uninfected counterparts. They had, for example, more prevalent diabetes (13.1% versus 7.9%), hypertension (39.2% versus 31.4%) and current cigarette smoking (29.3% versus 24.6%) (**Table 1**). Of note, matching was done on diabetes and hypertension as (yes/no) categories but not on HbA1c and systolic BP, respectively, as the latter (e.g., systolic BP) were included in the definition of the former (e.g., hypertension). Thus although matching did achieve overall balance in baseline traditional risk factor burden between the two exposure groups, as indicated by the composite measure of 30-year estimated ASCVD risk (SMD<0.01), participants with LTBI had marginally higher mean systolic BP (130 mmHg) than negative controls (127 mmHg; SMD=0.14).

### LTBI and all-cause mortality

After PS weight matching, participants with LTBI had an adjusted RMST (aRMST) (95%CI) at 18 years of 15.3 (14.8, 15.8) years while negative controls had an aRMST of 16.3 (15.8, 16.7) years (**Figure 2A-B**). Accordingly, LTBI was associated with a reduction in overall mean (95%CI) survival of 0.95 (0.26, 1.64) years (p=0.006) [*alternatively*, 11 (3, 29) months] at 18 years of follow-up (**Figure 2B**) relative to negative controls. Results for the unadjusted RMST analysis are presented in **Figure 2A**.

**Figure 2.**
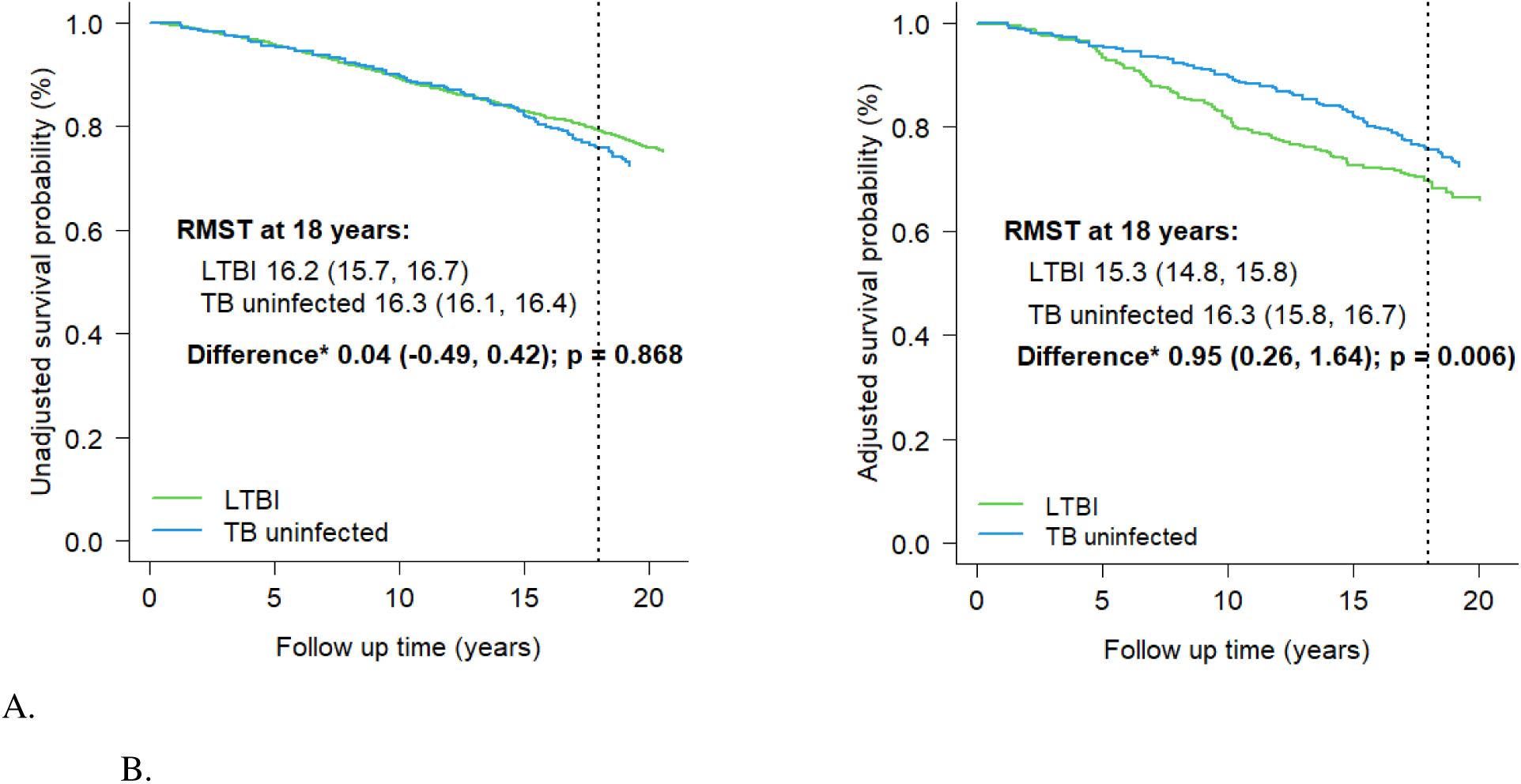
Kaplan-Meier curves for overall survival stratified by LTBI status (A) before and (B) after propensity-weighted matching, US NHANES 1999-2000 *Difference in RMST estimated as RMST for TB uninfected minus RMST for LTBI. Hazard ratio (HR) estimated in reference to LTBI group. aRMST = adjusted restricted mean survival time. Unadjusted = unmatched sample; adjusted = propensity weight matched sample.

### LTBI and cause-specific mortality

**Figures 3A-D** show the cumulative incidence functions and RMTL estimates at 18 years for cardiovascular and non-cardiovascular deaths in both unmatched and PS weight matched samples. We did not find evidence of differences in 18-year cardiovascular-specific mortality by LTBI status when accounting for competing risks in either unmatched [difference in RMTL (95%CI): 0.04 (−0.25, 0.33) years; p=0.771] (**Figure 3A**) or matched [difference in adjusted RMTL (aRMTL): 0.14 (−0.26, 0.54) years; p=0.483] (**Figure 3C**) analyses. We did, however, observe in matched analyses that LTBI [aRMTL: 2.01 (1.53, 2.49) years] was associated with greater lost life expectancy to non-cardiovascular death compared to being negative [aRMTL: 1.21 (0.84, 1.57) years] (**Figure 3D**). Alternatively, at 18 years follow-up, those with LTBI had lost (mean difference aRMTL) 0.81 (0.23, 1.43) years more than those negative to non-cardiovascular death (p=0.009).

**Figure 3.**
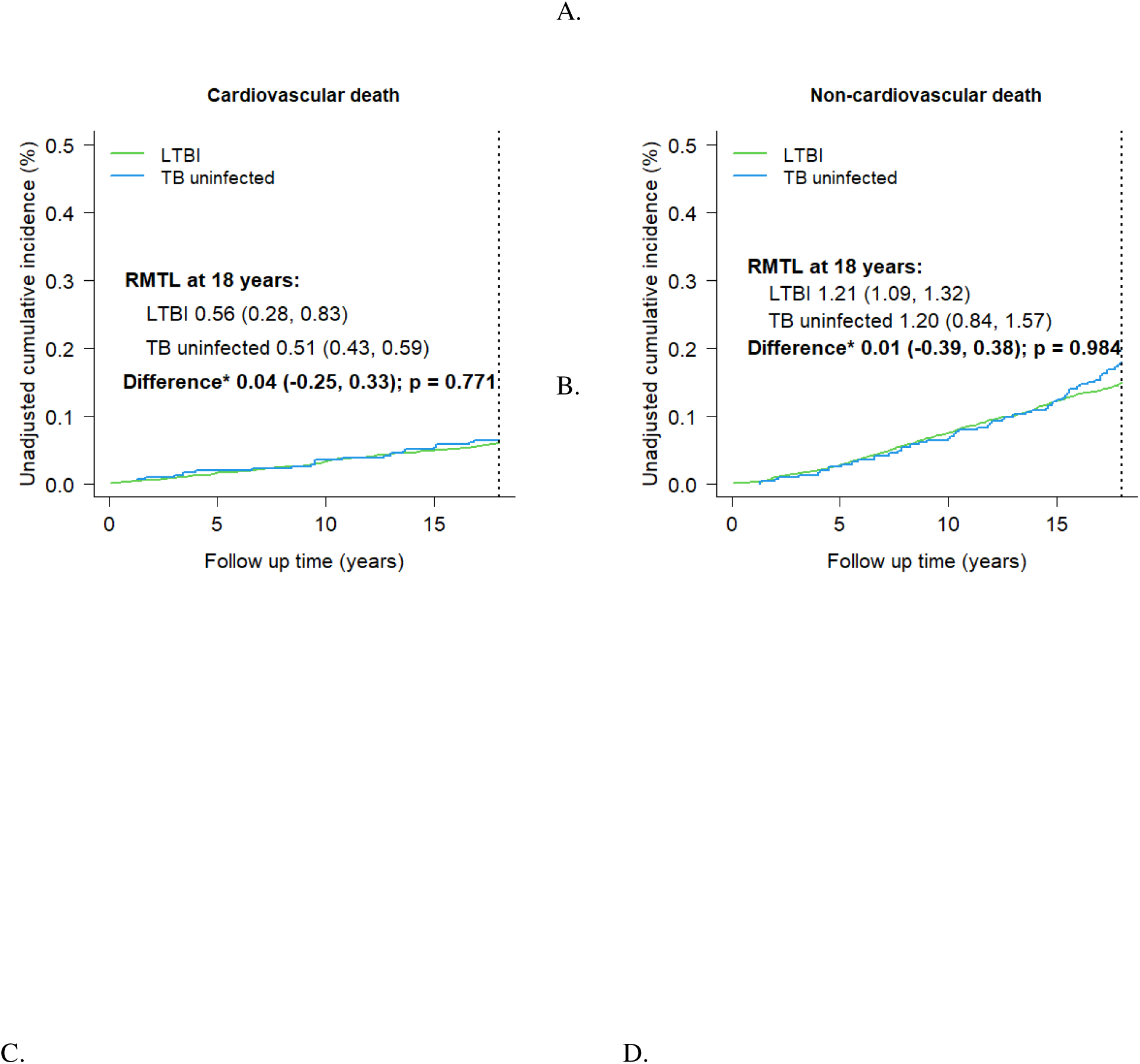
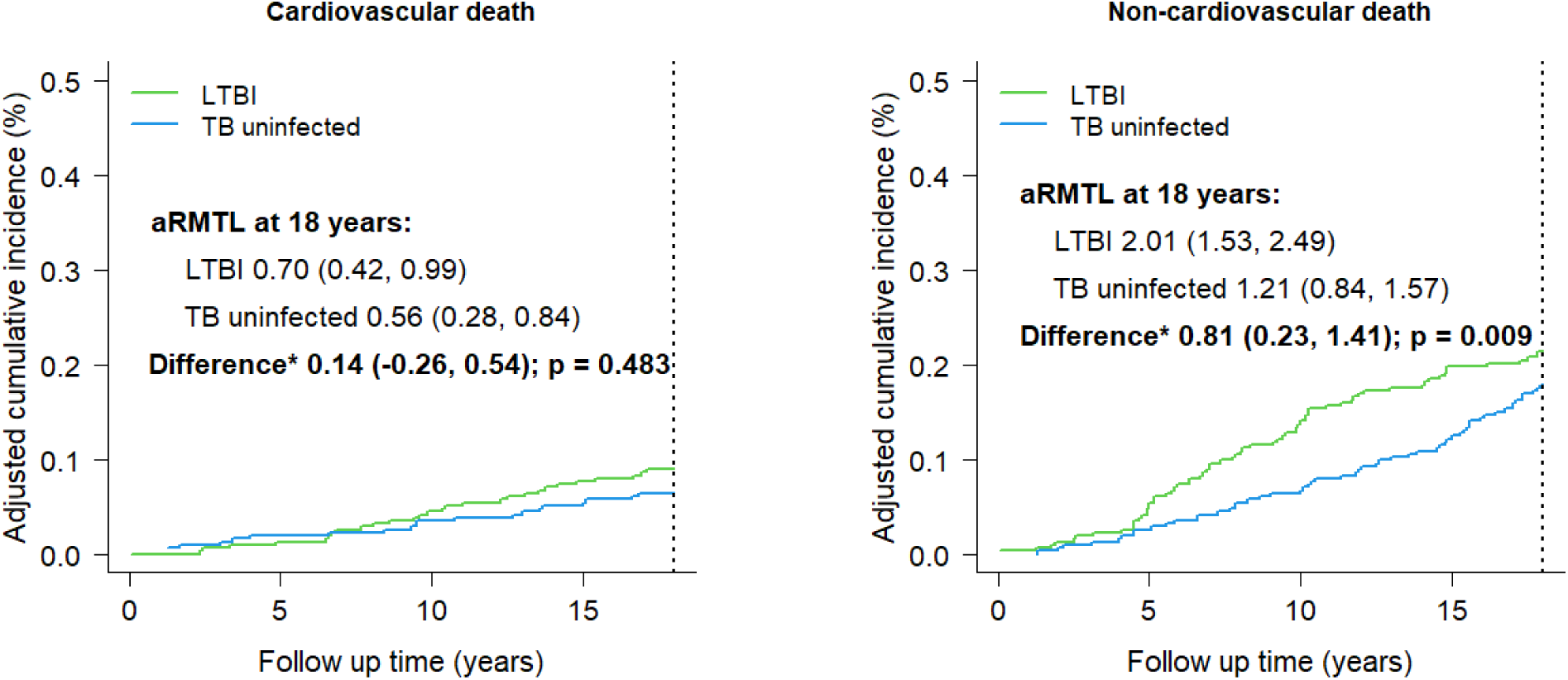
Cumulative incidence of cardiovascular and noncardiovascular cause-specific deaths during follow up stratified by stratified by latent tuberculosis infection status before and after propensity-weighted matching, US NHANES 1999-2000 *Difference in RMTL estimated as RMTL for TB uninfected minus RMTL for LTBI. aRMTL = adjusted restricted mean time lost. Unadjusted = unmatched sample; adjusted = propensity weight matched sample.

### Subgroup analyses

Statistically significant moderator effects were detected only for diabetes mellitus (p=0.004) (**Table 2**). Among non-diabetic participants, survival was comparable between those with LTBI [aRMST: 15.7 (15.2, 16.2) years] and negative controls [16.4 (15.9, 16.8)]. The aHR was 0.81 (0.59, 1.13) with LTBI as reference group. However, among those with diabetes at cohort entry, aRMST with LTBI was 11.5 (9.8, 13.3) years and 15.3 (14.0, 16.6) years without TB. This translated into nearly 4 years [3.8 (1.6, 6.0) years; p=0.001] shorter mean survival with co-prevalent LTBI and diabetes than diabetes alone. The aHR was 0.37 (0.21, 0.67) (p=0.007) with LTBI as reference group.

### Sensitivity Analysis

When LTBI was defined as skin induration ≥15mm, the matched sample was reduced to 139 participants with LTBI and another 139 negative controls (**Table 3**). There were 38 and 35 deaths with and without LTBI, respectively. Adjusted RMST was 16.0 (15.7, 16.4) years with LTBI and 16.7 (16.1, 17.3) years without, and thus a survival difference of 0.65 (−0.03, 1.33) years (p=0.058). In tandem with the primary analysis (skin induration ≥10mm), LTBI was associated with shorter overall survival [aRMST: 0.65 (−0.03, 1.33) years; p=0.058] compared to being negative. However, these differences disappeared at the lower threshold of skin induration ≥5mm. Adjusted RMST was 15.8 (15.4, 16.2) years with LTBI versus 16.0 (15.7, 16.4) years without. The corresponding difference in survival was 0.22 (−0.32, 0.97) years (p=0.141).

**Table 3.** Adjusted RMST for all-cause death by latent tuberculosis infection status and stratified by baseline characteristics, US NHANES 1999-2000.

| Subgroups | All-cause death |  |  |  |  |  |
| --- | --- | --- | --- | --- | --- | --- |
|  | Total deaths/Persons at-risk (n/N) |  | aRMST* at 18 years follow-up (years (95% CI)) |  |  |  |
|  | LTBI | Uninfected | LTBI | Uninfected | Difference | P value |
| *Primary analysis* |  |  |  |  |  |  |
| Overall: LTBI = induration $\geq 10$ mm | 96/313 | 86/313 | 15.3 (14.8, 15.8) | 16.3 (15.8, 16.7) | 0.95 (0.26, 1.64) | 0.006 |
| *Secondary analysis* |  |  |  |  |  |  |
| Overall: LTBI = induration $\geq 15$ mm | 38/139 | 35/139 | 16.0 (15.7, 16.4) | 16.7 (16.1, 17.3) | 0.65 (-0.03, 1.33) | 0.058 |
| Overall: LTBI = induration $\geq 5$ mm | 153/461 | 131/461 | 15.8 (15.4, 16.2) | 16.0 (15.7, 16.4) | 0.22 (-0.32, 0.97) | 0.141 |

\* Difference in RMST estimated as RMST for TB uninfected minus RMST for LTBI.
aRMST = adjusted restricted mean survival time; aHR = adjusted hazards ratio.
Adjusted by propensity-weighted matching.

## Discussion

This prospective study of US adults investigated the impact of LTBI on mortality, including from cardiovascular causes, and how such impact might vary according to baseline levels of traditional risk factors. We found that LTBI was associated with worse overall survival independent of demographic, socio-economic and traditional risk factors, among other potential determinants. This effect was particularly pronounced when LTBI was comorbid with diabetes mellitus. Findings were consistent in different sensitivity analyses. There were no significant differences in cardiovascular-specific survival by TST status, however. The study may have been underpowered to detect these. Future work should ascertain the causes and mechanisms of death in LTBI, while screening for traditional risk factors should be considered in precision targeting of LTBI preventive therapy.

LTBI is an otherwise asymptomatic condition. Whenever it has been in the crosshairs of clinical or population health research and practice, it has mainly been in relation to its potential progression to active TB, and the resultant individual morbidity and population transmission of *M.tb* (1), (2), (3), (40). Here we showed that LTBI is independently associated with worse overall survival. We unfortunately did not have data on specific underlying causes of death. We thus could not ascertain whether non-cardiovascular death related to subsequent active TB. Nonetheless, evidence shows that active disease is most likely to occur within the first two years following *M.tb* infection (41), (42). If no disease develops after 5 years the lifetime risk is estimated to be 2% and 0.5% after 10 years (41), (42), (43), (44). It has also been suggest that the majority of individuals thought to have LTBI, i.e., immune reactivity to TB antigens, have actually cleared their TB infection (45), (46). As such, the excess mortality in our study associated with LTBI over 18 years of follow up might not be due to progression to active TB. Conversely, the markedly diminished survival associated with co-occuring LTBI and diabetes might indicate how the latter increases the risk of both progressing to active TB(47) and experiencing adverse TB clinical outcomes, including mortality (48).

Although current systematic reviews and meta-analysis evidence suggests that persons with prior active TB may be at increased risk for incident cardiovascular events(11), the relationship between LTBI and cardiovascular health is less clear. Literature on the latter is sparse. The few available reports, drawn from Saudi Arabia (21), Egypt (22), Uganda (23)and Peru (23), (24) describe increased odds of prevalent atherosclerotic CVD (ASCVD) with LTBI. All these studies, however, are relatively small, hospital-based and either cross-sectional (21), (23) or case-control (22), (24). This lends them to criticism of potential survivorship and selection biases, temporal ambiguity, and limited external generalizability. To our knowledge, only one population-based prospective study to date has examined this relationship. Hossain *et. al.,* (2023) reported an 8% [aHR: 1.08 (0.99-1.18)] higher risk of incident ischemic heart disease or stroke with LTBI compared to no TB (25). Their study included 49,197 immigrants to Canada (1985– 2019) with total follow-up of 901,734 person-years and a median time of 19 years from cohort entry to censoring. We also found reduced cardiovascular-specific survival with LTBI in a mixed US cohort of immigrants and born citizens which was, however, not statistically significant.

It is noteworthy to highlight that our definition of LTBI based solely on TST, without complementary data on symptoms, chest imaging and/or sputum culture, subsumes a heterogenous group of tuberculous immunoreactive individuals (1), (3), (4), (5). This includes individuals with cleared *M.tb* infection, quiescent infection and active but subclinical infection (1), (4). These subgroups of tuberculous immunoreactivity likely signify divergent immunopathology (4), (5) and, in turn, differing cardiovascular and non-cardiovascular morbidity and mortality risk. Overlap with traditional risk factors may accentuate this risk as we observed with comorbid LBTI and diabetes mellitus in our results. From a mechanistic standpoint, an amplified and dysregulated inflammatory response to *M.tb* antigens drives insulin resistance, hypercoagulability, endothelial dysfunction and microvascular dysfunction, among others (18), (19), (49), (50), (51). Intermediate states like hypertension (52) and diabetes/hyperglycemia (53), (54), whose incidence is shown to be increased with LTBI, may be the proximate drivers of morbidity and mortality independent of progression to active TB.

### Strengths and limitations

Our study adds to the currently sparse literature on LTBI and its potential cardiovascular consequences. It also represents an invaluable perspective from a low TB incidence, high income setting. The results improve upon the currently available hospital-based reports with respect to generalizability. This is because NHANES samples are drawn to reflect the demographic, socio-economic and racial/ethnic diversity of the US population. However, the comparatively small analytic sample size post-matching reduced our statistical power to definitively detect cardiovascular-specific survival differences, if they exist. Relatedly, we lacked granular data on specific causes of death. Thus we could not account for the likely contribution of deaths from active TB. Our definition of LTBI based on TST alone risks misclassification of, for example, false positive results due to sensitization by environmental bacteria and BCG vaccination. Lastly, our primary analysis was based on RMST estimation, which is increasingly preferred to Cox proportional hazards modeling. RMST estimation is model-free while hazard ratio (HR) estimation depends on model assumptions that are rarely valid in practice. RMST estimates are absolute and can be easily comprehended. HR, on other hand, are relative estimates meaning the magnitude of absolute effects is unclear.

## Conclusion

LTBI may be associated with worse long-term overall survival, particularly when it co-occurs with diabetes. The specific mechanisms of death remain to be ascertained but may be other than cardiovascular or progression to active TB disease. This warrants further exploration especially in TB endemic regions of the world like sub-Saharan Africa and South-East Asia. Future work with more accurate definitions of LTBI should aim to definitively ascertain its impact on cardiovascular morbidity and mortality, while screening for co-occuring traditional risk factors should be considered in precision targeting of LTBI preventive therapy.

## Author contributions

IMM, KAW, RJW conceptualization; IMM and BC data curation, formal analysis and visualization; IMM, KAW, BC, AA, LM, MJS, MN, NABN, JMN, RJW investigation, methodology and data interpretation; IMM and AA project administration; IMM and RJW writing – original draft; and IMM, KAW, BC, AA, LM, MJS, MN, NABN, JMN, RJW writing – review & editing. All authors have read and approved the manuscript before submission.

## Data Availability

All data produced are available online at: https://www.cdc.gov/nchs/nhanes/about_nhanes.htm

## Acknowledgements

None.

## Online data supplemement

**Table E1.** ICD-10 codes used in cause-of-death categorization.

| **Cause-of-death category** | **ICD-10 codes** |
| --- | --- |
| Heart disease | I00 – I09, I11, I13, I20 – I51 |
| Cerebrovascular diseases | I60 – I69 |
| Cancer | C00 – C97 |
| Respiratory system diseases | J00 – J98 |
| Alzheimer’s and other dementias | G30, F00 – F09 |
| Digestive system diseases | K00 – K92 |
| Diabetes mellitus | E10 – E14 |
| Accidents (unintentional injuries) | V01 – X59, Y85 – Y86, 9998 |
| Urinary system diseases | N00 – N39 |
| Infectious and parasitic diseases | A00 – B99 |

